# Factors associated with unfavorable outcome after anterior circulation stroke despite successful reperfusion and early neurological improvement

**DOI:** 10.1101/2023.05.17.23290103

**Authors:** Jessica Jesser, Simon Nagel, Martin Bendszus, Silvia Schönenberger, Peter A. Ringleb, Christian Herweh, Markus A. Möhlenbruch, Johannes A. Vey, Thanh N. Nguyen, Charlotte S. Weyland

## Abstract

**Background and Purpose:** Successful reperfusion during endovascular therapy (EVT) usually leads to early neurological improvement (ENI), yet some patients do not achieve good long-term outcome. The aim of this study was to investigate factors associated with unfavorable clinical outcome (UCO) despite ENI.

**Methods:** This was a retrospective single-center analysis of consecutive patients treated for anterior circulation large vessel occlusion who achieved successful reperfusion and ENI (at least 20% lower follow-up NIHSS compared to NIHSS on admission) between 01/2014 and 04/2019. The primary endpoint was unfavorable outcome (90-day mRS > 2 or mRS > pre-stroke mRS). Patients with favorable clinical outcome (FCO) and UCO despite ENI were compared in univariable regression analysis.

**Results:** Successful EVT (mTICI 2c-3) and ENI occurred in 396/549 (72.1 %) patients and unfavorable outcome despite ENI in 168/396 (42.2 %) patients. Factors associated with UCO were pre-stroke mRS (Odds ratio (OR) 3.13 95% confidence interval (CI) 2.53–3.96, p < 0.001), age (OR 1.09 CI 1.07-1.12, p < 0.001), baseline NIHSs (OR 1.09 CI 1.06-1.13, p < 0.001), female sex (OR 1.92 CI 1.28-2.89, p = 0.002), hypertension (OR 2.32 CI 1.37-4.03, p = 0.002), smoking (OR 0.48 CI 0.25-0.87, p = 0.019), history of previous stroke (OR 2.7 CI 1.61-4.59, p < 0.001), atrial fibrillation (OR 1.73 CI 1.16-2.6, p = 0.007), neutrophil-lymphocyte ratio (OR 1.05 CI 1.01 – 1.09, p = 0.014), as well as ASPECTS after EVT (OR 0.77 CI 0.68-0.86, p < 0.001), TAN score (OR 0.60 CI 0.46-0.78, p < 0.001) and Swieten score (OR 2.55 CI 1.87-3.54, p < 0.001). Several ASPECTS regions were associated with UCO despite ENI (insula, M1-M3 and M5).

**Conclusions:** Relevant factors associated with UCO despite successful EVT and ENI were patient age, sex, pre-stroke mRS, hypertension, history of smoking, previous stroke, atrial fibrillation, neutrophil-lymphocyte ratio, TAN and Swieten Score as well as ASPECTS after EVT. The involvement of certain brain regions by ASPECTS segments were associated with UCO despite ENI after EVT.

## Introduction

Endovascular therapy (EVT) has changed the treatment paradigm for acute ischemic stroke (AIS) patients with increasing case numbers world-wide in the wake of demographic changes and widening of the indication for treatment [1]. Outcome prediction after ischemic stroke and EVT is important to aid patient and family counseling [2-5]. Successful revascularization of the target vessel occlusion is known to be pivotal for longterm favorable clinical outcome (FCO) [6, 7]. Nevertheless, recent meta-analyses showed that approximately half of the patients with successful EVT do not show favorable 90-day functional outcome (modified Rankin Scale 0-2) [4, 8] This has been referred to as futile recanalization [9, 10] or reperfusion without functional independence, as many patients can still have quality of life [8]. Understanding underlying mechanisms of reperfusion without functional independence (RFI) by applying prediction models may help to identify risks as well as protective factors, thus allowing for a more personalized therapy and potentially better recovery during stroke rehabilitation [11, 12].

Early neurological improvement (ENI) after successful EVT is known to be highly predictive for FCO [13, 14]. Predictors for ENI as well as predictors for delayed neurological improvement after AIS and EVT despite lack of ENI have been characterized [15]. However, in some patients, ENI after successful EVT in anterior circulation stroke with large vessel occlusion does not translate into long-term favorable clinical outcome. The aim of this study was to define factors associated with this phenomenon and to describe the association of long-term unfavorable clinical outcome (UCO) despite ENI as localized by specific ASPECTS brain regions affected by the stroke lesion.

## Methods

This was a retrospective single-center analysis of the (*>>blinded for peer-review<<)* consisting of prospectively treated patients with large vessel occlusion in the anterior circulation. The study was reported according to the Strengthening the Reporting of Observational Studies in Epidemiology (STROBE) criteria [16]. Patients treated between January 2014 and April 2019 with successful reperfusion (mTICI 2c or 3) were included. Treatment decisions were made according to local standard procedure following national or international guidelines. Between April 2014 and February 2016, the mode of sedation (conscious sedation vs. general anesthesia) was randomized within the SIESTA Trial [17]. Apart from SIESTA, we followed our local standard operating protocols. The preferred sedation modality was conscious sedation. Imaging protocol required at least either CT with CT-angiography or MRI with MR-Angiography [18]. Within (*>>blinded for peer-review<<)* all clinical parameters were obtained by NIHSS/mRS-certified neurologists. The clinical outcome as per mRS 90 days after stroke onset was obtained by a standardized interview (unblinded investigator by phone call or a personal letter to the patient). The recanalization result according to the modified Thrombolysis in Cerebral Infarction (mTICI) score was determined by the interventional neuroradiologist in charge of the EVT were included In the analysis. Intracranial hemorrhage (ICH) was defined as symptomatic according to the Heidelberg Bleeding Classification (HBC) [19].

### Primary endpoint and inclusion criteria

Patients with successful reperfusion, who experienced early neurological improvement (ENI), were included in further analyses (**Figure 1**). Early neurological improvement (ENI) was defined as a reduction of NIHSS by at least 20 % on discharge (or 3 days after admission) compared to NIHSS on admission (Kharitonova 2011, Stroke). The primary endpoint was unfavorable clinical outcome (UCO) (mRS > 2 or post-stroke mRS > pre-stroke mRS) 90 days after stroke onset despite ENI. Patients were grouped according to long-term clinical outcome into an UCO group and a favorable clinical outcome (FCO) group. Clinical, radiological and laboratory parameters were tested for the association with UCO despite successful EVT and ENI. These included demographics, pre-stroke co-morbidities (including diabetes mellitus type II, previous stroke, smoking, atrial fibrillation), initial NIHSS, pre-stroke modified Rankin Scale (mRS), time intervals to stroke therapy, sedation mode, intravenous thrombolysis using recombinant tissue plasminogen activator (rtPA); laboratory parameters on admission (neutrophil/lymphocyte ratio); location of vessel occlusion, tandem occlusions, admission ASPECTS, Tan-Score, Swieten scale, and presence of ICH after EVT (symptomatic or asymptomatic according to HBC).

**Figure 1.**
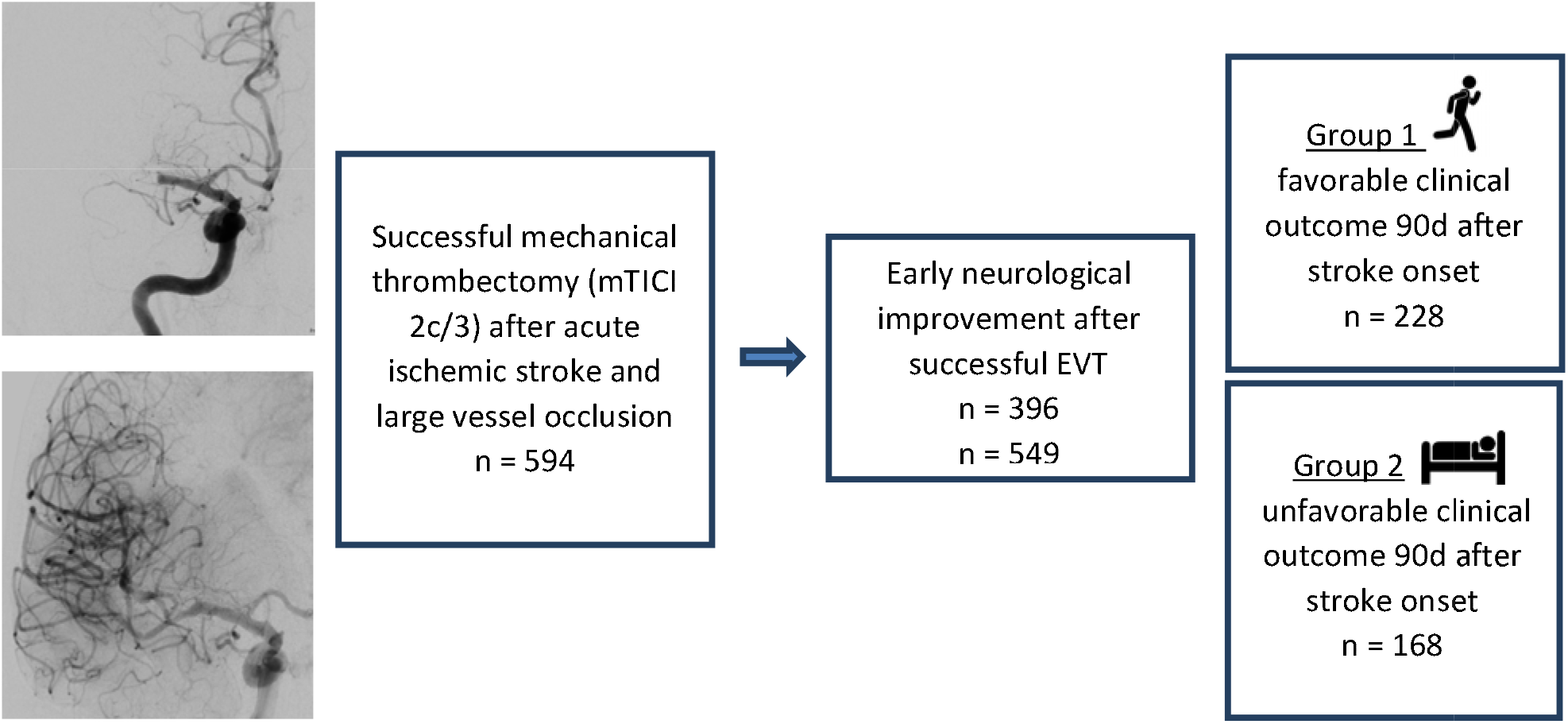
Patient selection and study.

The ASPECTS was automatically calculated (e-ASPECTS) on baseline CT-scan and verified by an experienced neuroradiologist (at least five years of radiological experience). [20, 21] The load of microangiopathic cerebral lesions according to the Swieten scale [22] and collateral status according to the Tan score [23] on baseline images were manually evaluated by an experienced neuroradiologist (at least five years of radiological experience). Follow-up imaging was routinely performed at our institution as cerebral CT-scan 20-36 hours after intervention and follow-up ASPECTS was also adjudicated by an experienced neuroradiologist (at least five years of radiological experience). Laboratory parameters were collected retrospectively from the electronic patient charts.

### Statistical analysis

The patient cohort was described regarding patients with favorable clinical outcome (FCO) vs. unfavorable clinical outcome (UCO) using summary measures of the empirical distribution. Continuous variables are presented as means (SD) and/or median (IQR and range), as appropriate. Nominal variables are described as absolute and relative frequencies. For descriptive analyses, continuous variables were tested using a *t*-test for independent groups, whereas categorical variables were compared applying the Chi²-test for unordered categorical variables or the U-test for ordered categorical variables. Univariable logistic regression was performed to examine the factors associated with UCO based on the complete cases, respectively. Furthermore, individual ASPECTS regions in follow-up imaging (CT 24 hours after intervention) irrespective of the affected hemisphere were analyzed conducting univariable and multivariable logistic regression modeling to assess their association with UCO. Since this was a retrospective data analysis, all p-values were interpreted in a descriptive sense; p-values < 0.05 were denoted as significant. All statistical analyses were performed using the software R version 4.0.2. (*>>blinded for peer-review<<)* and analysis within was approved by the local ethics committee (*>>blinded for peer-review<<)*. Patient consent was waived due to the retrospective character of this study.

## Results

During the study period, there were 594 patients who underwent EVT and achieved successful reperfusion (mTICI2c/3), of whom 396 had ENI. Of these, 168 (42%) had an unfavorable clinical outcome 90 days after stroke onset (**Figure 1**). Compared to patients, who had a favorable clinical outcome, patients with UCO were older (median [IQR], 80 [76–87] vs 69 [62–78], p < 0.001), had a higher median pre-stroke mRS (2 [1-3] vs. 0 [0–1], p < 0,001), were more likely to be female (62% vs. 46%, p < 0.001) and presented with a higher median NIHSS (17 [13-21] vs. 13 [8–18], p = < 0.001) (**Table 1**). They were more likely to have hypertension (87% vs. 74%, p = 0.002), atrial fibrillation (60% vs 47%, p=0.008), previous stroke (28% vs 12%, p = < 0.001) and were less likely to be smokers (10% vs 18%, p = 0.017) (**Table 1**). The median neutrophil-lymphocyte ratio in peripheral blood sampling after hospital admission was higher in patients with UCO (5.8 [4.2–9] vs. 4.5 [3.3– 6.7], p = 0.014) Patients with UCO had a comparable ASPECTS on admission (9 [8 – 10] for both study groups, p = 0.255), but a lower median ASPECTS after EVT (8 [6-9] vs 8 [7–10], p < 0.001). They also had a higher cerebral microangiopathy lesion load (median Swieten score: 2 [1-2] vs. 1 [1–1], p < 0.001) and a lower collateral score (median TAN score: 3 [2-3] vs. 3 [3-4], p < 0.001). The study groups did not differ concerning procedural aspects regarding time to treatment, vessel occlusion site, sedation mode, number of passes during thrombectomy, Table 1.

**Table 1:**
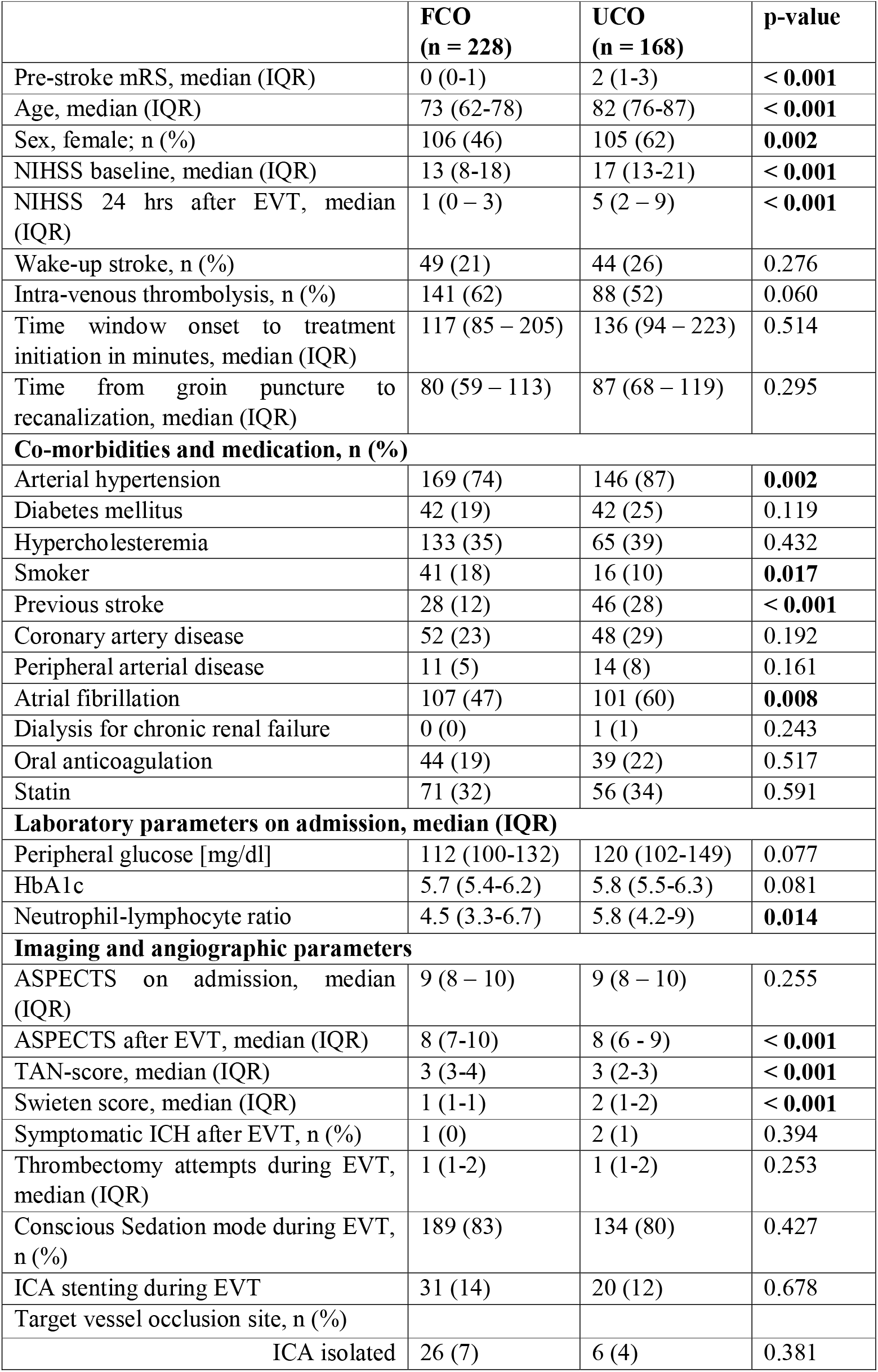
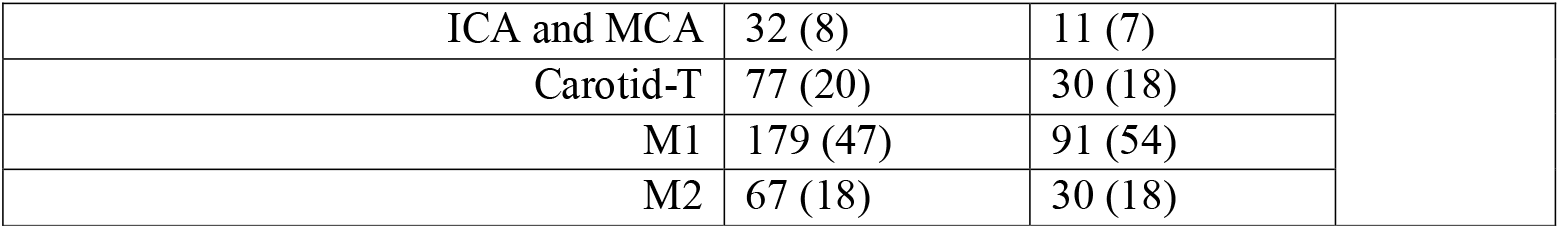
Baseline characteristics of patients with successful reperfusion and early neurological improvement with 90-day favorable clinical outcome (FCO) vs. unfavorable clinical outcome (UCO)

|  | **FCO**<br>**(n = 228)** | **UCO**<br>**(n = 168)** | **p-value** |
| --- | --- | --- | --- |
| Pre-stroke mRS, median (IQR) | 0 (0-1) | 2 (1-3) | **< 0.001** |
| Age, median (IQR) | 73 (62-78) | 82 (76-87) | **< 0.001** |
| Sex, female; n (%) | 106 (46) | 105 (62) | **0.002** |
| NIHSS baseline, median (IQR) | 13 (8-18) | 17 (13-21) | **< 0.001** |
| NIHSS 24 hrs after EVT, median (IQR) | 1 (0 – 3) | 5 (2 – 9) | **< 0.001** |
| Wake-up stroke, n (%) | 49 (21) | 44 (26) | 0.276 |
| Intra-venous thrombolysis, n (%) | 141 (62) | 88 (52) | 0.060 |
| Time window onset to treatment initiation in minutes, median (IQR) | 117 (85 – 205) | 136 (94 – 223) | 0.514 |
| Time from groin puncture to recanalization, median (IQR) | 80 (59 – 113) | 87 (68 – 119) | 0.295 |
| **Co-morbidities and medication, n (%)** |  |  |  |
| Arterial hypertension | 169 (74) | 146 (87) | **0.002** |
| Diabetes mellitus | 42 (19) | 42 (25) | 0.119 |
| Hypercholesteremia | 133 (35) | 65 (39) | 0.432 |
| Smoker | 41 (18) | 16 (10) | **0.017** |
| Previous stroke | 28 (12) | 46 (28) | **< 0.001** |
| Coronary artery disease | 52 (23) | 48 (29) | 0.192 |
| Peripheral arterial disease | 11 (5) | 14 (8) | 0.161 |
| Atrial fibrillation | 107 (47) | 101 (60) | **0.008** |
| Dialysis for chronic renal failure | 0 (0) | 1 (1) | 0.243 |
| Oral anticoagulation | 44 (19) | 39 (22) | 0.517 |
| Statin | 71 (32) | 56 (34) | 0.591 |
| **Laboratory parameters on admission, median (IQR)** |  |  |  |
| Peripheral glucose [mg/dl] | 112 (100-132) | 120 (102-149) | 0.077 |
| HbA1c | 5.7 (5.4-6.2) | 5.8 (5.5-6.3) | 0.081 |
| Neutrophil-lymphocyte ratio | 4.5 (3.3-6.7) | 5.8 (4.2-9) | **0.014** |
| **Imaging and angiographic parameters** |  |  |  |
| ASPECTS on admission, median (IQR) | 9 (8 – 10) | 9 (8 – 10) | 0.255 |
| ASPECTS after EVT, median (IQR) | 8 (7-10) | 8 (6 - 9) | **< 0.001** |
| TAN-score, median (IQR) | 3 (3-4) | 3 (2-3) | **< 0.001** |
| Swieten score, median (IQR) | 1 (1-1) | 2 (1-2) | **< 0.001** |
| Symptomatic ICH after EVT, n (%) | 1 (0) | 2 (1) | 0.394 |
| Thrombectomy attempts during EVT, median (IQR) | 1 (1-2) | 1 (1-2) | 0.253 |
| Conscious Sedation mode during EVT, n (%) | 189 (83) | 134 (80) | 0.427 |
| ICA stenting during EVT | 31 (14) | 20 (12) | 0.678 |
| Target vessel occlusion site, n (%) |  |  |  |
| ICA isolated | 26 (7) | 6 (4) | 0.381 |

|  |  |  |
| --- | --- | --- |
| ICA and MCA | 32 (8) | 11 (7) |
| Carotid-T | 77 (20) | 30 (18) |
| M1 | 179 (47) | 91 (54) |
| M2 | 67 (18) | 30 (18) |

Logistic regression analysis revealed the following factors significantly associated with UCO despite ENI after successful EVT: pre-stroke mRS (OR 3.13 CI 2.52 – 3.96, p < 0.001), patient age (OR 1.09 CI 1.07 - 1.12, p < 0.001), female sex (OR 1.92 CI 1.28 - 2.89, p = 0.002) arterial hypertension (OR 2.32 CI 1.37 – 4.03, p = 0.002), history of smoking (OR 0.48 CI 0.25 - 0.87, p < 0.019), atrial fibrillation (OR 1.73 CI 1.16 - 2.6, p = 0.008), neutrophil-lymphocyte ratio (OR 1.05 CI 1.01 - 1.09, p = 0.014) and ASPECTS after EVT (OR 0.77 CI 0.68 - 0.86, p < 0.001) (**Table 2**).

**Table 2.** Univariable logistic regression analysis. Factors associated with unfavorable clinical outcome (UCO) despite successful reperfusion and early neurological improvement (ENI) after anterior large vessel occlusion stroke

|  | **Univariable regression** |  |  |
| --- | --- | --- | --- |
|  | **Events/**<br>**patients**<br>**(missing**<br>**values)** | **OR (95% CI)** | **p-value** |
| **Pre-stroke mRS** | 167/395 (1) | 3.13 (2.52-3.96) | **< 0.001** |
| **Age** | 168/396 (0) | 1.09 (1.07-1.12) | **< 0.001** |
| **Sex female** | 168/396 (0) | 1.92 (1.28-2.89) | **0.002** |
| **Wake Up**<br>**Stroke** | 168/396 (0) | 1.30 (0.81-2.07) | 0.276 |
| **NIHSS baseline** | 168/396 (0) | 1.09 (1.06-1.13) | **< 0.001** |
| **Hypertension** | 168/396 (0) | 2.32 (1.37-4.03) | **0.002** |
| **Diabetes** | 168/396 (1) | 1.47 (0.9-2.39) | 0.120 |
| **Smoker** | 168/395 (8) | 0.48 (0.25-0.87) | **0.019** |
| **Previous Stroke** | 167/394 (2) | 2.70 (1.61-4.59) | **< 0.001** |
| **Atrial Fibrillation** | 167/395 (1) | 1.73 (1.16-2.6) | **0.008** |
| **NL ratio** | 153/369 (27) | 1.05 (1.01-1.09) | **0.014** |
| **ASPECTS baseline** | 162/381 (15) | 0.92 (0.79-1.06) | 0.248 |
| **ASPECTS follow-up** | 163/385 (11) | 0.77 (0.68-0.86) | **< 0.001** |
| **TAN score** | 131/304 (92) | 0.60 (0.46-0.78) | **< 0.001** |
| **Swieten score** | 160/380 (16) | 2.55 (1.87-3.54) | **< 0.001** |
| **Intravenous**<br>**thrombolysis** | 168/396 (0) | 0.68 (0.45-1.02) | 0.060 |
| **Thrombectomy passes** | 167/396 (3) | 1.09 (0.94-1.27) | 0.247 |
| **Sedation mode** | 168/396 (0) | 1.23 (0.74-2.05) | 0.427 |
| **Stent-assisted angioplasty** | 160/382 (14) | 0.88 (0.48-1.6) | 0.678 |
| **Time onset to treatment** | 123/303 (93) | 1.00 (1-1) | 0.526 |
| **Symptomatic ICH** | 168/396 (0) | 2.73 (0.26-59.14) | 0.413 |

When analyzing follow-up ASPECTS regions, ischemic involvement of the insula, M1-M5 regions were more often present in the UCO group (Table 3). In univariable logistic regression, ischemic involvement of the insula, M1-M3 and M5 regions were associated with UCO despite ENI (Table 4, Figure 2) disregarding the laterality (left vs. right hemisphere).

**Table 3.** Post-EVT Alberta Stroke Program Early CT Score (ASPECTS) regions. associated with unfavorable clinical outcome (UCO) despite early neurological improvement (ENI) after successful reperfusion. Follow-up imaging 24 to 72 hours after initial treatment, excluded for missing follow up imaging: 11 patients; p-value from Chi2 test.

| **Patients with follow-up imaging**<br>**(n = 385)** |  | **FCO**<br>**(n = 222)** | **UCO**<br>**(n = 163)** | **p-value** |
| --- | --- | --- | --- | --- |
|  | Caudate nucleus | 56 (25) | 43 (26) | 0.798 |
|  | Putamen | 81 (36) | 66 (40) | 0.424 |
|  | Internal capsule | 47 (21) | 42 (26) | 0.291 |
| **ASPECTS**<br>**regions**<br>**affected**<br>**by**<br>**ischemia,**<br>**n (%)** | Insula | 83 (37) | 86 (53) | **0.003** |
|  | M1 | 41 (18) | 46 (28) | **0.024** |
|  | M2 | 37 (17) | 58 (36) | **< 0.001** |
|  | M3 | 24 (11) | 31 (19) | **0.023** |
|  | M4 | 16 (7) | 20 (12) | 0.092 |
|  | M5 | 16 (7) | 22 (13) | **0.041** |
|  | M6 | 21 (9) | 22 (13) | 0.214 |

**Table 4.**
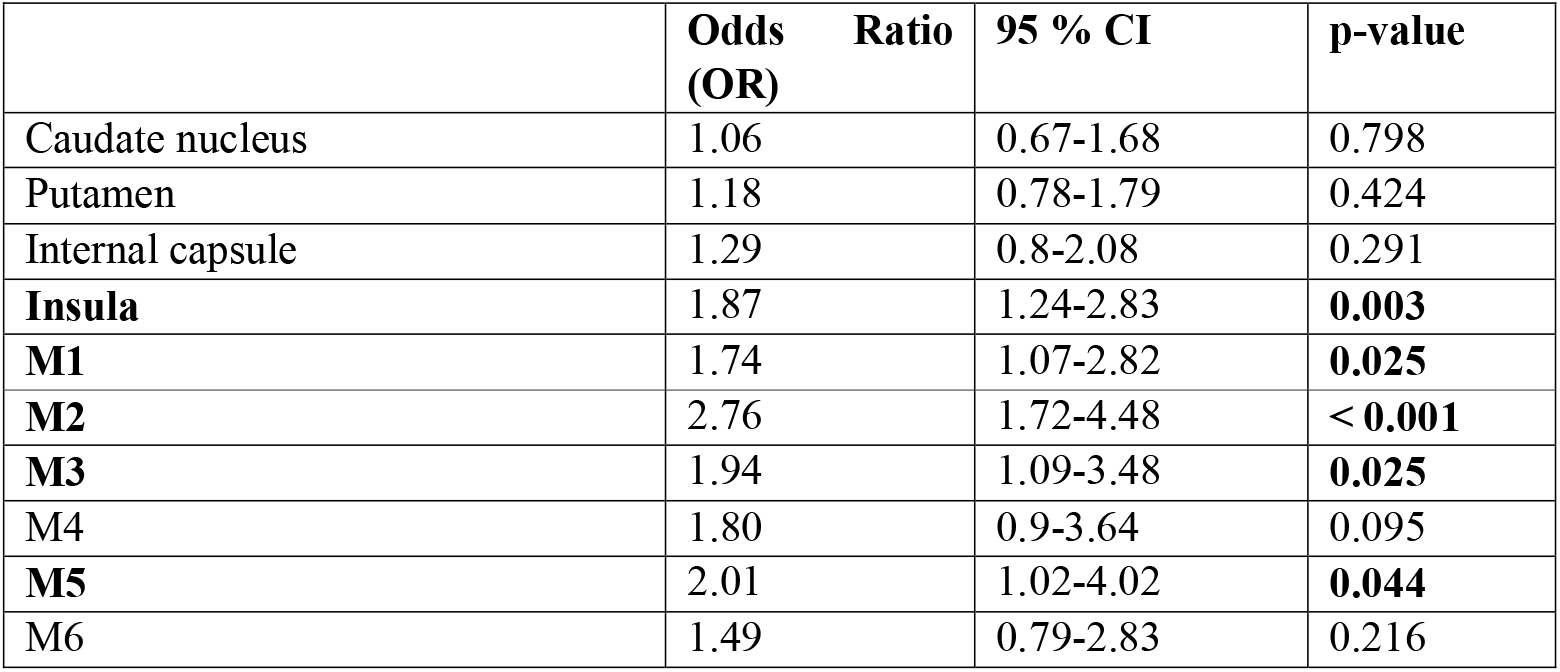
Univariable logistic regression for association of specific ASPECTS regions with unfavorable clinical outcome despite successful reperfusion and early neurological improvement. (patients excluded due to missing follow-up imaging = 11; follow-up CT or MRI 24 to 72 hours after initial therapy)

**Figure 2.**
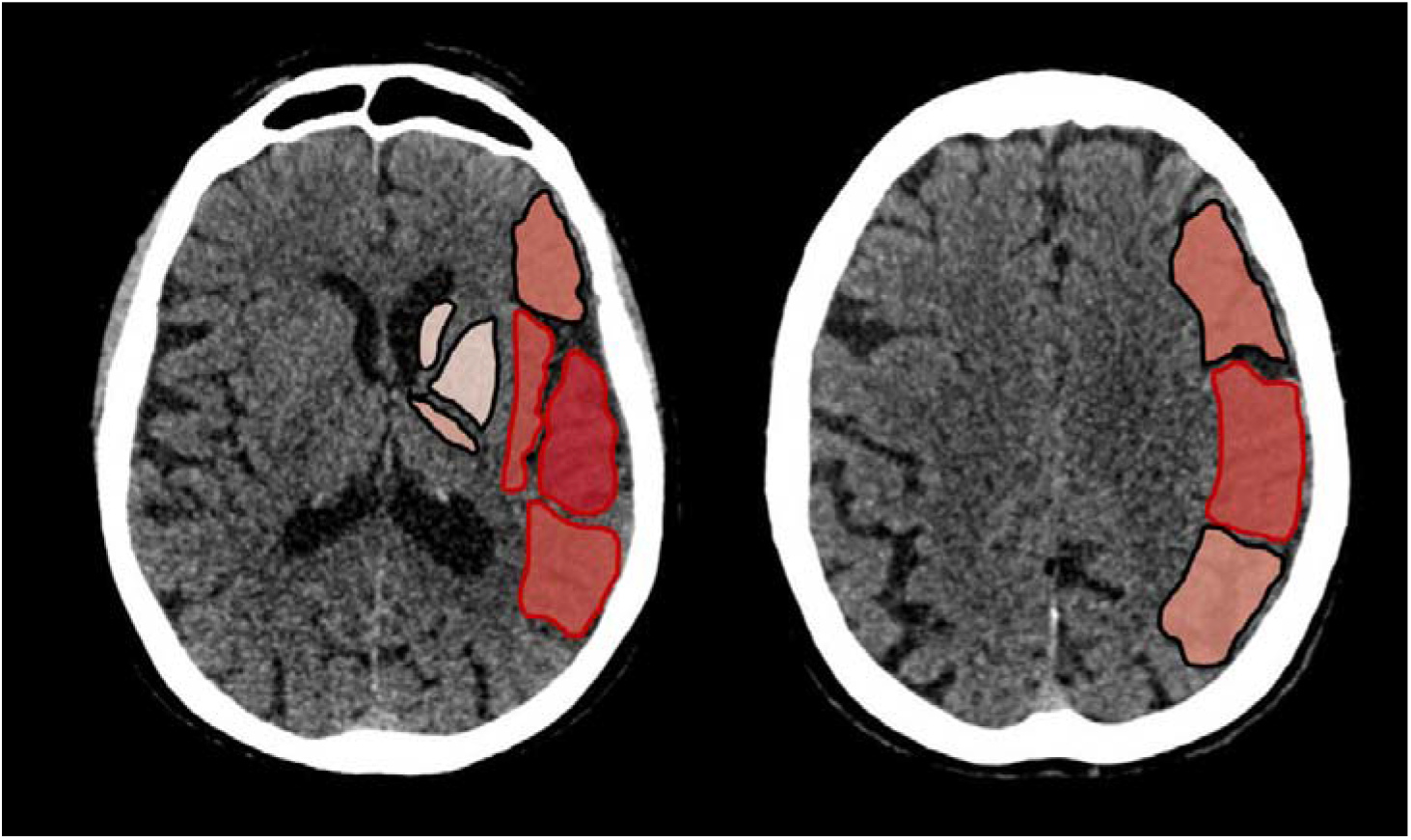
ASPECTS regions associated with UCO despite ENI. ASPECTS regions associated with unfavorable clinical outcome (UCO) despite early neurological improvement (ENI) outlined in red. The darker the color, the higher the odds of UCO for that region.

## Discussion

In this study of patients with anterior circulation large vessel occlusion undergoing EVT who achieved successful reperfusion and ENI, a substantial proportion of patients (42%) developed UCO at 90-day follow-up. This scenario of improving the neurological deficits in the days after the ischemic stroke event, but paradoxically not achieving good functional outcome can be due to different causes. Firstly, patients can be handicapped by stroke sequelae like pneumonia, seizures or even a second stroke event. Secondly, this paradoxon can be influenced by the indicative scores, namely NIHSS and mRS, that might measure different aspects of the patient’s clinical status.

Factors associated with 90-day UCO were pre-stroke disability, patient age, and baseline NIHSS, which are well known outcome predictors after AIS and EVT [24-26]. Our study showed that, even if patients achieved successful reperfusion and ENI, those who were more disabled, female, and older remained vulnerable to the development of later UCO. Moreover, vascular risk factors including hypertension, atrial fibrillation, and a medical history of a previous stroke were correlated with UCO despite ENI, whereas a history of smoking was found to be protective from UCO despite ENI.

Successful reperfusion after EVT and ENI are two endpoints known to be correlated with 90-day favorable clinical outcome. It is possible that early success achieved with these two benchmarks could breed complacency in the aftermath of patient rehabilitation. An understanding of factors associated with UCO despite successful reperfusion may help identify subgroups of patients who may merit close monitoring and targeted therapy with ENI after EVT. Female sex was associated with UCO despite ENI in our study group. There is emerging evidence about existing sex-related difference in stroke epidemiology and outcome but the related causes remain largely unknown [27]. Women are often older than men at the time of their index stroke and have a longer delay to hospital admission but despite these differences, women can still be as likely to achieve a good clinical outcome after ischemic stroke and endovascular stroke treatment [28]. Nonetheless, caregiver or community factors may also play a role in a patient’s recovery after stroke. In accordance with our results, an analysis from the SELECT study showed that despite men and women having similar discharge outcomes after anterior circulation EVT, women had worse functional outcome 90 days after stroke compared to men suggesting a potential role of post-discharge factors [29].

Paradoxically, a history of smoking was associated with better clinical outcome after stroke, as has been shown [30], which might reflect the observation that smokers have better intracranial vessel collaterals due to the chronic vascular stress and might therefore have developed tolerance to vessel occlusions [31]. UCO despite ENI was also correlated with a low collateral status (evaluated by the TAN score) [32], more white matter brain damage (Swieten Scale) [33], and larger stroke lesion in follow-up imaging (ASPECTS score FU) [34], as would be expected. Our study also showed the neutrophil-to-lymphocyte ratio (NLR) to be associated with UCO despite ENI. The NLR emerged recently as prognostic biomarker for clinical outcome after large vessel occlusion stroke [35-37] and might identify a target group who may benefit from adjunctive anti-inflammatory therapies. Despite promising results in experimental studies, inflammation-modulating treatments have not yet been translated successfully into the clinical setting [38].

With regards to imaging biomarkers, the association of infarct volume and clinical outcome after AIS is unclear, as some studies report final infarct volume as an independent predictor for outcome after stroke [39], whereas others do not [40]. This discrepancy underscores the notion that the relationship between final infarct volume and clinical outcome may be non-linear. Our study showed that lesions in certain brain areas were more susceptible to UCO despite ENI after successful EVT. Among these regions was the insula, which has been reported to be related to poor clinical outcome after stroke [41]. It is thought that the impairment of the insula’s role in the modulation of the autonomic nervous system leads to increased metabolic stress levels and may hinder rehabilitation after stroke. Furthermore, regions involved in motor control such as the M5 ASPECTS region and regions involved in language processing (M1-M3) were related to UCO despite ENI in our study. Among the affected regions, the strongest effect was shown by ASPECTS region M2, which is the temporal opercular region, which in most people is involved in sensory language processing. Lesions in this region are linked to a patient’s hindered potential for language rehabilitation as studies could show that patients with lesions in this area show global long-term aphasia which can affect their reintegration into daily life [42]. When comparing our results to other studies, one should bear in mind, that we analyzed a subgroup of patients who initially showed ENI. One study found that failed ENI was linked to lesions in the internal capsule as well as M4 and M5 regions. When the corticospinal tract in the internal capsule is harmed substantially it has been shown that motor function improvement is nearly impossible. Whereas cortical regions in the M5 regions, which are represented in the study group showing no ENI as well as in our study group showing ENI but UCO, might have the potential for some early neurological improvement dependent on the exact lesion location and extent but the rehabilitation potential might be limited in the long run [43]. Another study found similar results to our study, showing that involvement of the caudate, the insula and the M4 region were associated with poor long-term outcome [44].

Stratifying patients according to infarct locations associated with a greater risk of causing long-term disability might influence endovascular treatment strategies and call for individualized and intensified rehabilitation programs in some scenarios.

Limitations of this study are related to the single-center retrospective method, limiting generalizability to a broader population. The findings in follow-up imaging can vary depending on the imaging modality used (MRI vs. CT). The local standard operating procedures, post-procedure care can influence treatment strategies and study results. The substantial cohort size did allow for univariate regression analysis pointing out associated factors, that should be further evaluated. However, the amount of analyzed variables and clinical variables’ cross-influence reduced feasibility of multivariable logistic regression, so that this study cannot define independent predictors for UCO despite ENI. When analyzing the effect of ASPECTS areas on the clinical outcome of patients we did not take into account stroke laterality so as not to reduce statistical power of the analysis. Nonetheless, we acknowledge that hemispheric dominance might have an influence on patient outcomes and we suggest that similar analyses should be conducted on larger patient samples also accounting for stroke laterality.

## Conclusion

Unfavorable clinical outcome despite ENI after successful EVT is a common occurrence. Factors associated with delayed neurological deterioration, meaning unfavorable outcome despite successful endovascular therapy and early neurological improvement, were patient age, female sex, pre-stroke disability, arterial hypertension, smoking, history of stroke, neutrophil-lymphocyte ratio, TAN and Swieten Score, atrial fibrillation as well as baseline NIHSS and post-EVT ASPECTS. The ischemic involvement of the insular cortex, as well as other cortical ASPECTS brain regions, especially the M2 region, but also M1, M3 and M5, were associated with reperfusion without functional independence despite ENI.

## Data Availability

All data produced in the present work are contained in the manuscript

## Non-standard Abbreviations and Acronyms

ASPECTS: Alberta stroke program Early CT Score
CV: cross-validation
ENI: early neurological improvement
EVT: Endovascular therapy
FCO: favorable clinical outcome
HBC: Heidelberg Bleeding Classification
ICH: intracranial hemorrhage
LVO: large vessel occlusion
mRS: modified Rankin Scale
mTICI: modified Treatment In Cerebral Ischemia
NIHSS: National Institutes of Health Stroke Scale
rtPA: recombinant tissue Plasminogen Activator
RFI: reperfusion without functional independence
UCO: Unfavorable clinical outcome

## References

1. Ragoschke-Schumm, A. and S. Walter, DAWN and DEFUSE-3 trials: is time still important? Radiologe, 2018. 58(Suppl 1): p. 20–23.

2. Matsumoto, K., et al., Stroke Prognostic Scores and Data-Driven Prediction of Clinical Outcomes After Acute Ischemic Stroke. Stroke, 2020. 51(5): p. 1477–1483.

3. Goyal, M., et al., Challenges of Outcome Prediction for Acute Stroke Treatment Decisions. Stroke, 2021. 52(5): p. 1921–1928.

4. Weyland, C.S., et al., Full Reperfusion Without Functional Independence After Mechanical Thrombectomy in the Anterior Circulation : Performance of Prediction Models Before Versus After Treatment Initiation. Clin Neuroradiol, 2022.

5. Brugnara, G., et al., Multimodal Predictive Modeling of Endovascular Treatment Outcome for Acute Ischemic Stroke Using Machine-Learning. Stroke, 2020. 51(12): p. 3541–3551.

6. Tonetti, D.A., et al., Successful reperfusion, rather than number of passes, predicts clinical outcome after mechanical thrombectomy. J Neurointerv Surg, 2019.

7. Ospel, J.M., et al., Clinical impact of EVT with failed reperfusion in patients with acute ischemic stroke: results from the ESCAPE and ESCAPE-NA1 trials. Neuroradiology, 2021. 63(11): p. 1883–1889.

8. Seker, F., et al., Reperfusion Without Functional Independence in Late Presentation of Stroke With Large Vessel Occlusion. Stroke, 2022. 53(12): p. 3594–3604.

9. Goyal, M., et al., Endovascular thrombectomy after large-vessel ischaemic stroke: a meta-analysis of individual patient data from five randomised trials. Lancet, 2016. 387(10029): p. 1723–31.

10. Deng, G., et al., Predictors of futile recanalization after endovascular treatment in acute ischemic stroke: a meta-analysis. J Neurointerv Surg, 2022. 14(9): p. 881–885.

11. Nie, X., et al., Clinically Ineffective Reperfusion After Endovascular Therapy in Acute Ischemic Stroke. Stroke, 2023. 54(3): p. 873–881.

12. Neuberger, U., et al., Optimal thresholds to predict long-term outcome after complete endovascular recanalization in acute anterior ischemic stroke. J Neurointerv Surg, 2021. 13(12): p. 1124–1127.

13. Weyland, C.S., et al., Predictors for Failure of Early Neurological Improvement After Successful Thrombectomy in the Anterior Circulation. Stroke, 2021. 52(4): p. 1291–1298.

14. Soize, S., et al., Can early neurological improvement after mechanical thrombectomy be used as a surrogate for final stroke outcome? J Neurointerv Surg, 2019. 11(5): p. 450–454.

15. Talavera, B., et al., Delayed Neurological Improvement After Full Endovascular Reperfusion in Acute Anterior Circulation Ischemic Stroke. Stroke, 2021. 52(7): p. 2210–2217.

16. von Elm, E., et al., The Strengthening the Reporting of Observational Studies in Epidemiology (STROBE) statement: guidelines for reporting observational studies. Lancet, 2007. 370(9596): p. 1453–7.

17. Schönenberger, S., et al., Effect of Conscious Sedation vs General Anesthesia on Early Neurological Improvement Among Patients With Ischemic Stroke Undergoing Endovascular Thrombectomy: A Randomized Clinical Trial. JAMA, 2016. 316(19): p. 1986–1996.

18. Nagel, S., et al., Simplified selection criteria for patients with longer or unknown time to treatment predict good outcome after mechanical thrombectomy. J Neurointerv Surg, 2019. 11(6): p. 559–562.

19. von Kummer, R., et al., The Heidelberg Bleeding Classification: Classification of Bleeding Events After Ischemic Stroke and Reperfusion Therapy. Stroke, 2015. 46(10): p. 2981–6.

20. Nagel, S., et al., e-ASPECTS software is non-inferior to neuroradiologists in applying the ASPECT score to computed tomography scans of acute ischemic stroke patients. Int J Stroke, 2017. 12(6): p. 615–622.

21. Nagel, S., et al., Clinical Utility of Electronic Alberta Stroke Program Early Computed Tomography Score Software in the ENCHANTED Trial Database. Stroke, 2018. 49(6): p. 1407–1411.

22. van Swieten, J.C., et al., Grading white matter lesions on CT and MRI: a simple scale. J Neurol Neurosurg Psychiatry, 1990. 53(12): p. 1080–3.

23. Tan, I.Y.L., et al., CT Angiography Clot Burden Score and Collateral Score: Correlation with Clinical and Radiologic Outcomes in Acute Middle Cerebral Artery Infarct. American Journal of Neuroradiology, 2009. 30(3): p. 525.

24. Singer, O.C., et al., Age dependency of successful recanalization in anterior circulation stroke: the ENDOSTROKE study. Cerebrovasc Dis, 2013. 36(5-6): p. 437–45.

25. Sharobeam, A., et al., Functional Outcomes at 90 Days in Octogenarians Undergoing Thrombectomy for Acute Ischemic Stroke: A Prospective Cohort Study and Meta-Analysis. Front Neurol, 2019. 10: p. 254.

26. Siegler, J.E., et al., Endovascular vs Medical Management for Late Anterior Large Vessel Occlusion With Prestroke Disability: Analysis of CLEAR and RESCUE-Japan. Neurology, 2023. 100(7): p. e751–e763.

27. Rexrode, K.M., et al., The Impact of Sex and Gender on Stroke. Circ Res, 2022. 130(4): p. 512–528.

28. Sun, D., et al., Sex-Related Differences in Outcomes of Endovascular Treatment for Anterior Circulation Large Vessel Occlusion. Stroke, 2023. 54(2): p. 327–336.

29. Fifi, J.T., et al., Sex differences in endovascular thrombectomy outcomes in large vessel occlusion: a propensity-matched analysis from the SELECT study. J Neurointerv Surg, 2023. 15(2): p. 105–112.

30. von Martial, R., et al., Impact of smoking on stroke outcome after endovascular treatment. PLoS One, 2018. 13(5): p. e0194652.

31. Vagal, A., et al., Collateral Clock Is More Important Than Time Clock for Tissue Fate. Stroke, 2018. 49(9): p. 2102–2107.

32. Nambiar, V., et al., CTA collateral status and response to recanalization in patients with acute ischemic stroke. AJNR Am J Neuroradiol, 2014. 35(5): p. 884–90.

33. Mistry, E.A., et al., White Matter Disease and Outcomes of Mechanical Thrombectomy for Acute Ischemic Stroke. AJNR Am J Neuroradiol, 2020. 41(4): p. 639–644.

34. van Horn, N., et al., Predictors of poor clinical outcome despite complete reperfusion in acute ischemic stroke patients. J Neurointerv Surg, 2021. 13(1): p. 14–18.

35. Goyal, N., et al., Admission Neutrophil-to-Lymphocyte Ratio as a Prognostic Biomarker of Outcomes in Large Vessel Occlusion Strokes. Stroke, 2018. 49(8): p. 1985–1987.

36. Wang, L., et al., Neutrophil to lymphocyte ratio predicts poor outcomes after acute ischemic stroke: A cohort study and systematic review. J Neurol Sci, 2019. 406: p. 116445.

37. Paudel, S.S., B. Thapa, and R. Luitel, Neutrophil Lymphocyte Ratio as a Prognostic Marker in Acute Ischemic Stroke: a Systematic Review and Meta-analysis. J Nepal Health Res Counc, 2021. 18(4): p. 573–579.

38. Drieu, A., et al., Anti-inflammatory treatments for stroke: from bench to bedside. Ther Adv Neurol Disord, 2018. 11:p. 1756286418789854.

39. Boers, A.M.M., et al., Association of follow-up infarct volume with functional outcome in acute ischemic stroke: a pooled analysis of seven randomized trials. J Neurointerv Surg, 2018. 10(12): p. 1137–1142.

40. Johnston, K.C., et al., Combined clinical and imaging information as an early stroke outcome measure. Stroke, 2002. 33(2): p. 466–72.

41. Timpone, V.M., et al., Percentage insula ribbon infarction of >50% identifies patients likely to have poor clinical outcome despite small DWI infarct volume. AJNR Am J Neuroradiol, 2015. 36(1): p. 40–5.

42. Plowman, E., B. Hentz, and C. Ellis, Jr., Post-stroke aphasia prognosis: a review of patient-related and stroke-related factors. J Eval Clin Pract, 2012. 18(3): p. 689–94.

43. Weyland, C.S., et al., Predictors for Failure of Early Neurological Improvement After Successful Thrombectomy in the Anterior Circulation. Stroke, 2021. 52(4): p. 1291–1298.

44. Seyedsaadat, S.M., et al., Differential Contribution of ASPECTS Regions to Clinical Outcome after Thrombectomy for Acute Ischemic Stroke. AJNR Am J Neuroradiol, 2021. 42(6): p. 1104–1108.

